# Burden of Neglected Tropical Diseases in Ethiopia: Analysis from the Global Burden of Disease Estimates 2023

**DOI:** 10.64898/2026.01.19.26344412

**Authors:** Jemal Beksisa, Alemu Earsido, Ally Walker, Asnake Worku, Awoke Misganaw, Chalie Mulu, Haftu Reda, Jonathan F Mosser, Mohsen Naghavi, Shikur Mohammed, Sebsibe Tadesse, Tamrat Mekonnen, Tigist Shumet, Wondimye Ashenafi, Yenework Acham, Zenabu Reda, Simon I Hay

## Abstract

**Introduction:** Neglected Tropical Diseases (NTDs) comprises over 20 preventable infectious illness that disproportionately affecting low- and middle-income countries, including Ethiopia. Although national control initiatives have expanded since 2013, evidence on recent national and subnational burden trends remains limited. This study aimed to assess trends in mortality and disability-adjusted life years attributable to neglected tropical diseases in Ethiopia at national and subnational level from 1990 to 2023.

**Methods:** We used data from the Global Burden of Disease 2023 study to estimate age-standardized and age-specific mortality and Disability-Adjusted Life Years (DALYs) attributable to neglected tropical diseases. Results are presented as absolute numbers and age-standardized rates (per 100,000 population), disaggregated by disease, sex, region, and year from 1990 to 2023, with 95% uncertainty intervals.

**Result:** In 2023, the national age-standardized DALY rate for all NTDs was 708.3 where, schistosomiasis accounted the largest share (228.7; 95% UI: 145.4–375.0), followed by other NTDs (104.4; 95% UI: 59.7–171.4), trachoma (100.0; 95% UI: 67.4–144.9), and rabies (78.3; 95% UI: 9.5–257.9), together contributed to over three-quarters DALYs due to NTDs. The national age-standardized mortality rate was 5.98 mostly contributed by schistosomiasis (2.01; 95% UI: 1.72–2.38) and rabies (1.40; 95% UI: 0.18–4.58). Substantial regional variation was observed, with Addis Ababa recording the lowest (200.5 and 1.2) and Afar the highest (860.3 and 8.9) DALYs and mortality rate due to all NTDs respectively. Between 1990 and 2023, DALYs and mortality rate declined by 87.5% and 91.6%, respectively, although declines slowed after 2010. During 2010–2023, increases in DALYs and mortality due to schistosomiasis were observed in Addis Ababa (+25%) and Harari (+13%), while trachoma increased in Oromia (+9%).

**Conclusion:** NTDs continue to impose a substantial burden of disability and mortality in Ethiopia, with pronounced regional disparities and a slowing pace of progress in recent years. The observed stagnation or increases in specific NTDs at the subnational level highlight the need for sustained monitoring and targeted control strategies.

## Introduction

Neglected tropical diseases are a group of bacterial, parasitic and viral infections that predominantly affect populations in tropical and sub-tropical regions, particularly where poverty is rampant. These diseases are termed “neglected” because they receive limited attention in global health agenda, attract insufficient funding, and disproportionately affect poor and marginalized communities, leading to stigma, disability, and social exclusion(1). NTDs are grouped together due to their chronic and often disabling effects, their strong association with poverty, and their overlapping geographic distribution. The term “neglected tropical diseases” was first introduced by the World Health Organization (WHO) in 2007 to unify disease control initiatives under a single coordinated global program. In 2012, WHO classified 17 diseases as NTDs, and by 2017 this list expanded to 20, forming the current global NTD portfolio (2–4).

Globally, NTDs are among the top ten communicable causes of ill health, contributing relatively low mortality but substantial morbidity (5). In 2021, WHO estimated that more than one billion people were infected with at least one neglected tropical disease (NTD), approximately 1.5 billion additional people were at risk, and NTDs accounted for an estimated 120,000 deaths and 14.1 million disability-adjusted life years (DALYs) globally each year(6,7). Infection may lead to blindness, disfigurement, severe disability, malnutrition, or impaired childhood development, and individuals are often infected with multiple diseases simultaneously. Although, NTDs are endemic to nearly 150 countries, the greatest burden is concentrated in low- and middle-income countries in Africa, Asia, and Latin America. Rural populations and urban slum residents are disproportionately affected due to inadequate access to clean water, sanitation, health care, and improved housing (8,9).

The African Region bears approximately 40% of the global NTD burden, with all 47 countries in the region endemic for at least one NTD, and 36 of them (78%) co-endemic for at least five or more diseases(10–12). In Ethiopia, communicable diseases, including NTDs, continue to pose a major public health challenges in the Sustainable Development Goals (SDG) era(13–16). The national NTDs mapping performed in 2013 showed that around 80 million people live in areas endemic to at least one NTD, and nearly all regions are affected by at least three or more diseases, with the highest burden concentrated in central, western and northwestern parts of the country(17,18).

The Global efforts to combat NTDs have been shaped by WHO roadmaps, beginning with the 2012–2020 and continuing with the current roadmap, “Ending the Neglect to Attain the SDGs: 2021–2030,” which targets prevention, control, elimination, or eradication of 20 NTDs. This roadmaps outline five core strategies implemented by countries: preventive chemotherapy through mass drug administration (MDA) or the SAFE strategy; intensified disease management; vector and intermediate host control; veterinary public health interventions for zoonotic NTDs; and improved water, sanitation, and hygiene (WASH) to interrupt transmission(4,19–21). In alignment with these frameworks, Ethiopia, completed national NTD mapping and launched its first NTD master plan in 2013, followed by strategic plans for 2013–2015, 2016–2020, and 2021–2025(18,22,23). The first two plans targeted nine priority diseases: trachoma, onchocerciasis, schistosomiasis, soil-transmitted helminths, lymphatic filariasis, podoconiosis, leishmaniasis, dracunculiasis, and scabies, while the third plan expanded to include rabies, leprosy, and dengue or chikungunya. Across all strategic periods, Ethiopia has implemented the 5 WHO-recommended intervention pillars.

Ethiopia has implemented programs to control and eliminate NTDs over the past decades; however, the overall trend in disease burden whether increasing or decreasing remains unclear. Although third national the third national strategic plan (2021–2025) is nearing completion, the impact of these nationwide efforts on mortality and disability remains poorly quantified. Existing studies on NTD burden in Ethiopia are limited in scope, mostly localized, and lack comprehensive national or subnational estimates(13,17,24,25). Evidence on long-term trends in NTD-related mortality and DALYs over the past decades remains scarce. In this context, the Global Burden of Disease 2023 study provides the most reliable and comparable population-level estimates for assessing disease burden at both national and subnational levels.

Therefore, this study aims to estimate national and subnational mortality and DALY rates, as well as their trends, for NTDs in Ethiopia from 1990 to 2023. The findings will provide essential evidence on the burden of NTDs and the impact of past and ongoing control interventions, supporting policymakers and program leaders in strengthening future prevention and control strategies to reduce the national NTD burden.

## Materials and Methods

### Study setting

Ethiopia is the second most populous country in Africa after Nigeria and ranks tenth globally. According to the World Population Prospects (2023), the country’s total estimated population was 128,691,692 with an annual growth rate of 2.5%, a fertility rate of 3.88 and the median age of 18.7 years. The country is administratively divided into twelve regional states and two city administrations. Over 77% of the population resides in rural areas where access to basic health care and sanitation facilities are limited(26). This analysis reported Central and South Ethiopian region under South Nation Nationalities and Peoples Regional State(SNNP)

### Data source

This study used the most recent GBD 2023 estimates. The GBD provides systematically modeled measures of disease burden for all countries and subnational administrative levels, disaggregated by age and sex. For NTDs, the GBD synthesizes multiple data sources, including mortality and disease surveillance systems, routine health information reports, WHO databases, outbreak notifications, national surveys, and peer-reviewed publications. For Ethiopia, additional inputs include data from the Federal Ministry of Health surveillance platforms and national program reports. All data sources undergo standardization, bias adjustment, and quality assessment prior to modeling. Comprehensive documentation of GBD data sources and analytical protocols is publicly available through the GBD data repository(27,28).

### Case Definition and Cause list

In GBD 2023, neglected tropical diseases are defined as communicable diseases caused by parasitic, bacterial, viral, and other pathogens that disproportionately affect populations in tropical and subtropical settings and result in substantial morbidity, disability, and premature mortality. NTDs are classified under the Level 3 cause hierarchy. The cause list includes schistosomiasis, Soil-Transmitted Helminthiases (STH), onchocerciasis, lymphatic filariasis, leishmaniasis (visceral and cutaneous), African trypanosomiasis, yellow fever, Ebola, zika virus, Guinee worm, Chagas disease, dengue, rabies, trachoma, cysticercosis, echinococcosis, food-borne trematodiases, leprosy, and other NTDs not modeled individually. The “other NTDs” category comprises bacterial, viral, and parasitic infections such as relapsing fevers, Lyme disease, typhus, tick-borne spotted fevers, arboviral diseases, viral hemorrhagic fevers, cestode infections, and other helminth and protozoal infections. These conditions are mapped to multiple International Classifications of Disease (ICD-10) (A68–A98, B33–B83, P37) and ICD-9 (065–129.0) codes to capture their contributions to mortality and morbidity.

### Estimation of Mortalities and DALYs

GBD 2023 estimated both crude and age-standardized mortality rates for NTDs by age, sex, region, and year. Cause-specific mortality estimation methods varied by disease and data availability and included Cause of Death Ensemble Models (CODEm), custom count-based regression models applied to standardized cause-of-death data, and natural history models that derive mortality from modeled incidence and case-fatality rates. Non-fatal outcomes were estimated using a range of approaches, including DisMod-MR, a Bayesian hierarchical compartmental meta-regression model, spatiotemporal Gaussian process regression, model-based geostatistics, and other disease-specific modeling strategies. The selection of modeling approaches was guided by epidemiological characteristics and data availability for each NTD. Disability-adjusted life years (DALYs) were calculated as the sum of years of life lost (YLLs) due to premature mortality and years lived with disability (YLDs). YLLs were computed by multiplying cause-specific deaths by the standard life expectancy at the age of death, while YLDs were estimated by multiplying disease prevalence by corresponding disability weights. Uncertainty was propagated throughout the modeling process, and all estimates are reported with 95% uncertainty intervals derived from 1,000 posterior draws. Full details on GBD data and methods are available on the GBD website” and the GBD 2023 fatal and non-fatal capstone papers(29,30).

### Trend Analysis

Temporal trends in NTD-related mortality and DALYs were assessed using annual percent change (APC), calculated from age-standardized rates and reported with 95% uncertainty intervals. Trends were examined for the periods 1990–2023, 2000–2010, and 2010–2023, consistent with standard GBD analytical practice. National and regional estimates for Ethiopia were extracted by sex and age group to assess long-term trends and recent changes in NTD burden.

### Ethics Statement

This analysis was conducted as part of the GBD Collaborator Network and follows the established GBD research protocol (IHME ID: 4239-GBD2019-042022). The GBD study uses secondary, de-identified, and aggregated data sources. As such, informed consent is not required. Ethical approval, including a waiver of informed consent, was reviewed and granted by the Institutional Review Board of the University of Washington. Detailed information on GBD ethical approvals is publicly available through the GBD research documentation(28).

## Result

### National and Subnational Disability Adjusted Life Years due to NTDs, 2023

In 2023, neglected tropical diseases, combined with malaria, accounted for an estimated 1,802,606 (95% UI: 825,566–3,225,938) number of DALYs in Ethiopia. NTDs alone contributed 730,787 DALYs, representing 40.5% of the combined NTD–malaria burden and approximately 1.4% of DALYs from all causes. Of the total NTD DALYs, 359,548 (49.2%) occurred among males and 371,239 (50.8%) among females.

At the national level, the age-standardized DALY rate (ASDALY) for all NTDs in 2023 was 708.26 per 100,000 population, with slightly higher rates among females (732 per 100,000) than males (684 per 100,000). Schistosomiasis was the leading contributor to NTD-related ASDALYs, with 228.70 per 100,000 population (95% UI: 145.37–374.99), followed by other NTDs (104.35; 95% UI: 59.67–171.36), trachoma (99.96; 95% UI: 67.35–144.90), and rabies (78.32; 95% UI: 9.48–257.90). Together, these four disease groups accounted for approximately three-quarters of the national NTD DALY burden, with schistosomiasis alone contributing 32.4% (Table 1).

**Table 1.**
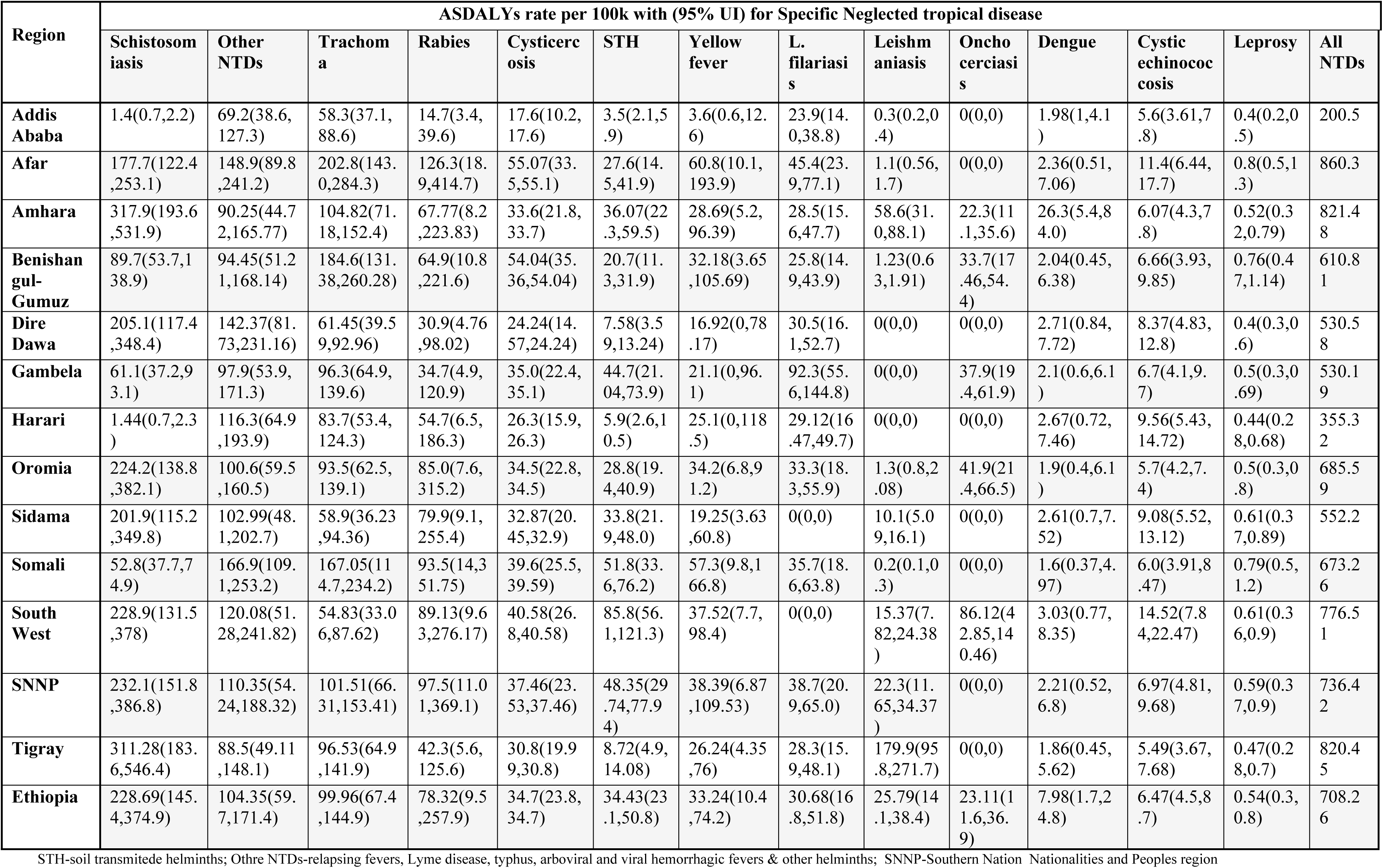
National and Subnational Age standardized DALYs Rate Due to NTDs 2023.

Across most regions, schistosomiasis was the predominant contributor to NTD-related DALYs, followed by trachoma and other NTDs. Subnational ASDALY rates varied substantially, ranging from 200.5 per 100,000 population in Addis Ababa to 860.3 per 100,000 in Afar Region (Figure 1).

**Figure 1.**
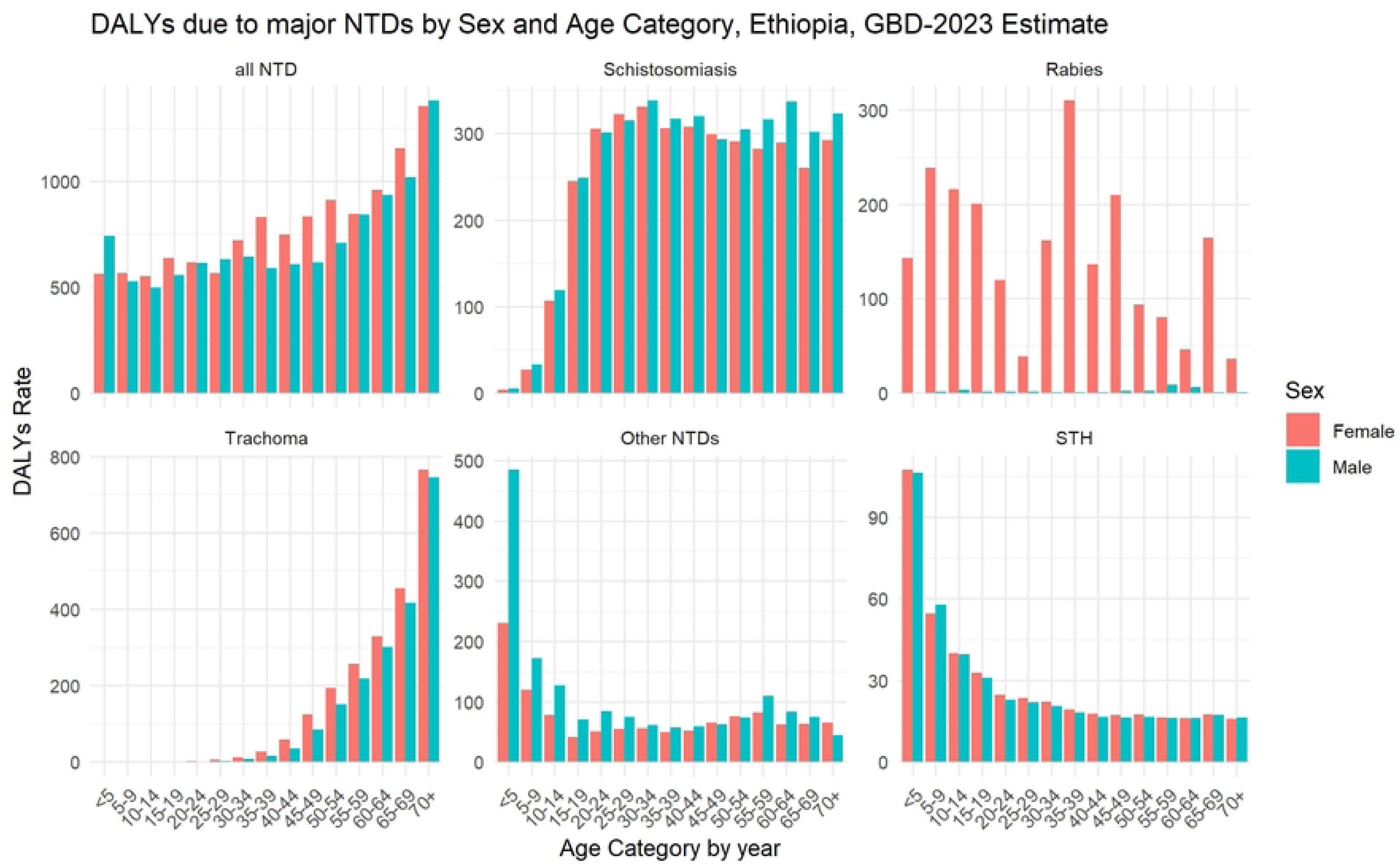
Subnational distirbution of ASDALYs and ASMR due to all neglected tropical diseases in 2023, Ethiopia.

#### Age- and sex-specific patterns of DALYs due to neglected tropical diseases

Overall NTD DALYs increased with age, with the highest burden observed among individuals aged 60 years and older. For most major NTDs; schistosomiasis, trachoma, soil-transmitted helminth (STH) infections, and other NTDs, ASDALY rates were broadly similar between males and females. In contrast, rabies showed a pronounced sex difference, with a substantially higher ASDALY rate among females (155.63 per 100,000; 95% UI: 14.87–516.97) compared with males (1.76 per 100,000; 95% UI: 0.15–5.14) (Figure 2).

**Figure 2.**
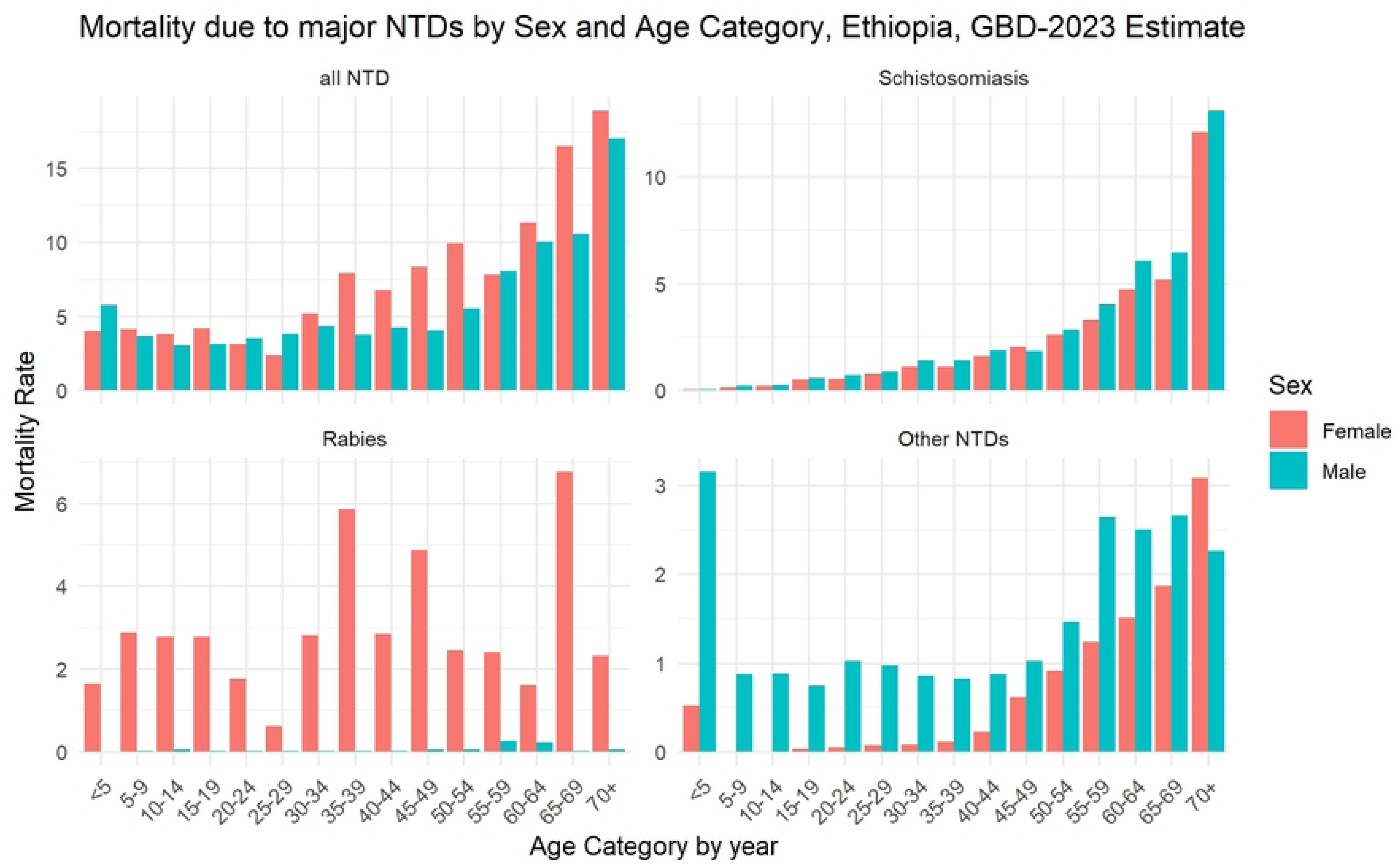
Age- and sex-specific DALY rates for major neglected tropical diseases in Ethiopia, GBD 2023.

Age-specific patterns differed by disease. STH infections and other NTDs predominantly affected younger populations, particularly children under 15 years of age. Trachoma showed a marked shift toward older age groups, with the highest burden among individuals aged 50 years and above. Schistosomiasis and rabies exhibited relatively consistent DALY burdens across age groups (Figure 2).

### National and Subnational Mortality due to Neglected Tropical Disease, 2023

In 2023, NTDs combined with malaria were responsible for an estimated 17,700 deaths (95% UI: 6,568–33,556) in Ethiopia. Of these, 5,467 deaths were attributable to NTDs alone, accounting for 30.8% of deaths from the combined NTD–malaria burden and approximately 0.7% of all-cause mortality. Among NTD-related deaths, 2,897 (52.9%) occurred among females and 2,570 (47.1%) among males.

The national age-standardized mortality rate (ASMR) for all NTDs was 5.98 per 100,000 population, with higher rates among females (6.50 per 100,000) than males (5.45 per 100,000). Schistosomiasis was the leading cause of NTD-related mortality, with an ASMR of 2.01 per 100,000 (95% UI: 1.72–2.38), followed by rabies (1.40; 95% UI: 0.18–4.58), other NTDs (0.97; 95% UI: 0.39–1.94), and yellow fever (0.54; 95% UI: 0.17–1.21). These four causes together accounted for over 82% of NTD-related deaths nationally (Table 2).

**Table 2.** National and Sub national Age Standardized Mortality Rate due to NTDs, 2023.

| Region | ASMR per 100k with (95% UI) for Specific Neglected tropical disease |  |  |  |  |  |  |  |  |  |
| --- | --- | --- | --- | --- | --- | --- | --- | --- | --- | --- |
|  | Schistosomiasis | Rabies | Other NTDs | Yellow fever | Leishmaniasis | Cysticercosis | Dengue | STH | Cystic echinococcosis | All NTDs |
| **Addis Ababa** | 0.04(0.02,0.07) | 0.26(0.06,0.69) | 0.66(0.23,1.50) | 0.06(0.01,0.20) | 0(0.00,0.00) | 0.06(0.03,0.11) | 0.02(0.01,0.05) | 0.02(0.01,0.03) | 0.09(0.05,0.13) | 1.21 |
| **Afar** | 3.53(2.16,5.06) | 2.26(0.37,7.09) | 1.2(0.47,2.45) | 0.97(0.17,3.12) | 0.02(0.01,0.02) | 0.42(0.22,0.68) | 0.05(0.01,0.17) | 0.28(0.13,0.45) | 0.17(0.09,0.26) | 8.9 |
| **Amhara** | 2.41(1.82,3.12) | 1.21(0.15,3.94) | 1.1(0.39,2.27) | 0.46(0.08,1.55) | 0.8(0.41,1.20) | 0.3(0.17,0.47) | 0.69(0.14,2.23) | 0.08(0.06,0.11) | 0.09(0.06,0.12) | 7.15 |
| **Benishangul-Gumuz** | 1.67(0.88,2.63) | 1.14(0.20,3.87) | 0.98(0.39,1.97) | 0.51(0.06,1.70) | 0.02(0.01,0.03) | 0.52(0.31,0.81) | 0.05(0.01,0.15) | 0.17(0.08,0.28) | 0.09(0.05,0.13) | 5.14 |
| **Dire Dawa** | 1.81(0.89,2.96) | 0.55(0.09,1.68) | 0.93(0.34,2.00) | 0.27(0.00,1.20) | 0(0.00,0.00) | 0.15(0.07,0.24) | 0.05(0.01,0.18) | 0.05(0.01,0.11) | 0.14(0.08,0.21) | 3.95 |
| **Gambela** | 1.09(0.57,1.88) | 0.62(0.10,2.13) | 0.96(0.37,2.04) | 0.34(0.00,1.53) | 0(0.00,0.00) | 0.34(0.18,0.56) | 0.04(0.01,0.13) | 0.17(0.05,0.34) | 0.1(0.06,0.16) | 3.67 |
| **Harari** | 0.04(0.02,0.07) | 1(0.13,3.43) | 1.01(0.38,2.13) | 0.41(0.00,1.94) | 0(0.00,0.00) | 0.15(0.07,0.25) | 0.06(0.02,0.18) | 0.05(0.02,0.09) | 0.16(0.09,0.25) | 2.87 |
| **Oromia** | 2.02(1.55,2.55) | 1.56(0.14,5.72) | 0.86(0.38,1.62) | 0.56(0.11,1.45) | 0(0.00,0.00) | 0.28(0.17,0.45) | 0.04(0.01,0.15) | 0.11(0.08,0.14) | 0.08(0.05,0.11) | 5.51 |
| **Sidama** | 1.45(0.84,2.09) | 1.41(0.19,4.45) | 1.18(0.44,2.46) | 0.3(0.06,0.94) | 0.14(0.07,0.22) | 0.25(0.15,0.38) | 0.06(0.01,0.17) | 0.25(0.14,0.38) | 0.14(0.08,0.20) | 5.17 |
| **Somali** | 0.94(0.62,1.27) | 1.67(0.26,6.26) | 0.89(0.36,1.78) | 0.93(0.16,2.80) | 0(0.00,0.00) | 0.21(0.12,0.34) | 0.04(0.01,0.12) | 0.32(0.22,0.47) | 0.08(0.04,0.11) | 5.08 |
| **South West** | 2.16(1.21,3.13) | 1.53(0.19,4.66) | 1.42(0.50,3.08) | 0.59(0.12,1.55) | 0.21(0.10,0.33) | 0.4(0.23,0.62) | 0.06(0.02,0.19) | 0.73(0.46,1.10) | 0.23(0.13,0.35) | 7.32 |
| **SNNP** | 2.25(1.75,2.88) | 1.76(0.21,6.66) | 1.07(0.42,2.14) | 0.59(0.11,1.67) | 0.31(0.16,0.47) | 0.26(0.14,0.41) | 0.05(0.01,0.15) | 0.16(0.11,0.21) | 0.1(0.06,0.14) | 6.54 |
| **Tigray** | 2.05(1.51,2.73) | 0.77(0.11,2.24) | 0.85(0.33,1.68) | 0.43(0.07,1.21) | 2.48(1.32,3.72) | 0.2(0.12,0.30) | 0.04(0.01,0.13) | 0.04(0.02,0.07) | 0.08(0.05,0.11) | 6.92 |
| **Ethiopia** | 2.01(1.73,2.37) | 1.4(0.18,4.59) | 0.97(0.39,1.94) | 0.53(0.17,1.21) | 0.36(0.19,0.53) | 0.27(0.16,0.41) | 0.21(0.05,0.65) | 0.15(0.11,0.18) | 0.09(0.06,0.13) | 5.98 |

STH-soil transittede helminths; Othre NTDs-relapsing fevers, Lyme disease, typhus, arboviral and viral hemorrhagic fevers & other helminths; SNNP-Southern Nation Nationalities and Peoples region

Mortality increased with age for most NTDs, peaking among individuals aged 60 years and older. Schistosomiasis and other NTDs showed similar age and sex patterns, with progressively increasing mortality across age groups. Rabies mortality was consistently higher among females (2.78 per 100,000; 95% UI: 0.27–9.17) compared with males (0.04 per 100,000; 95% UI: 0.002–0.12) and was highest among adult age groups rather than children or the elderly (Figure 3).

**Figure 3.**
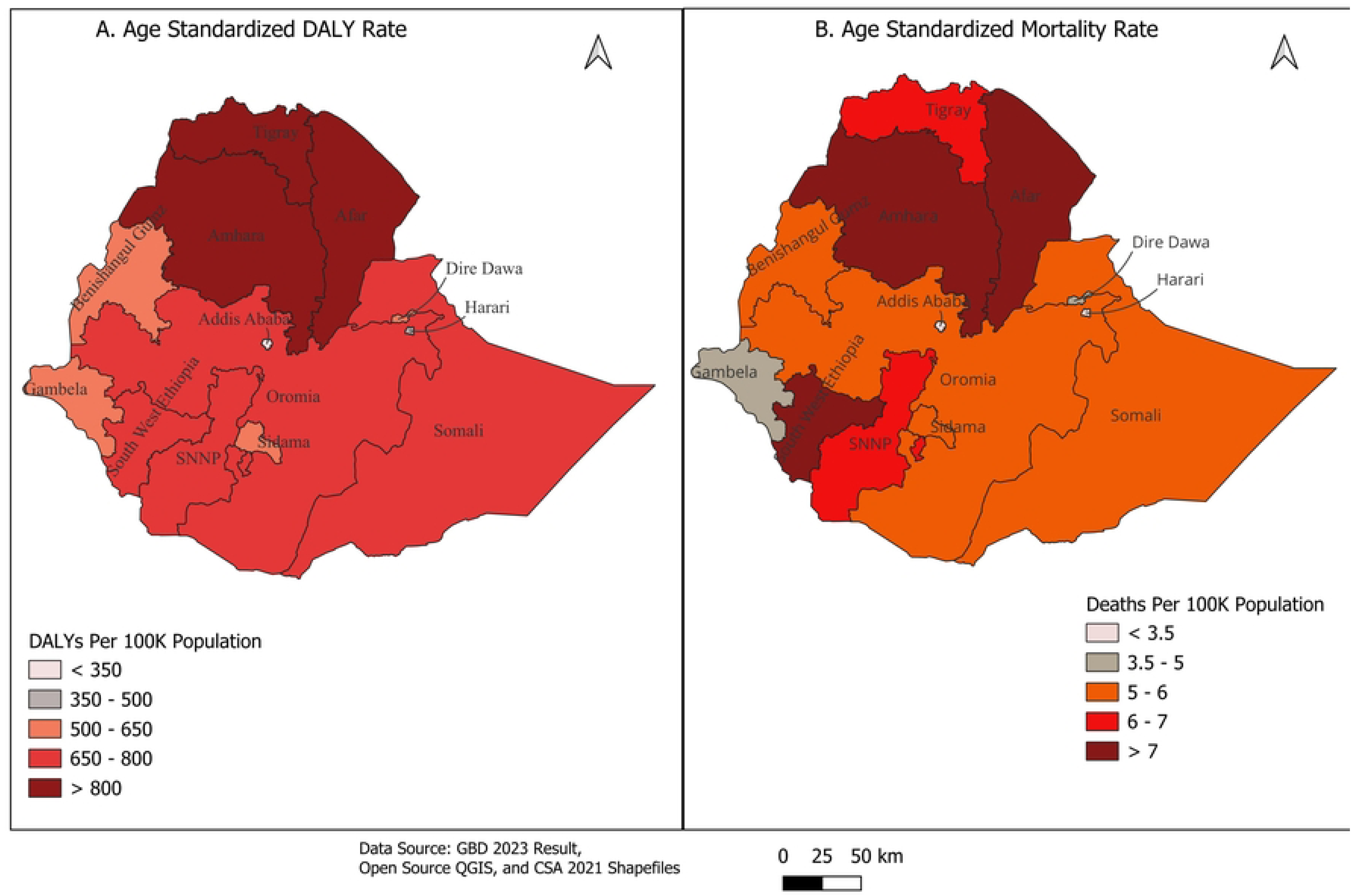
Age- and sex-specific Death rates for major neglected tropical diseases in Ethiopia, GBD 2023.

Sub nationally, schistosomiasis and rabies were the leading causes of NTD-related mortality in most regions. However, leishmaniasis was the primary cause in Tigray, while other NTDs were the leading contributors in Addis Ababa. Overall ASMRs ranged from 1.21 per 100,000 in Addis Ababa to 8.90 per 100,000 in Afar Region (Figure 1).

### National and Subnational trends in DALYs and Mortality due to NTDs, 1990-2023

Between 1990 and 2023, the national ASDALY rate for all NTDs declined from 5,658.9 per 100,000 to 708.3 per 100,000, representing an overall reduction of 87.5%. However, the magnitude of reduction slowed over time, with a 50.5% decrease during 2000–2010 compared with a 37.0% decrease during 2010–2023 (Figure 4).

**Figure 4.**
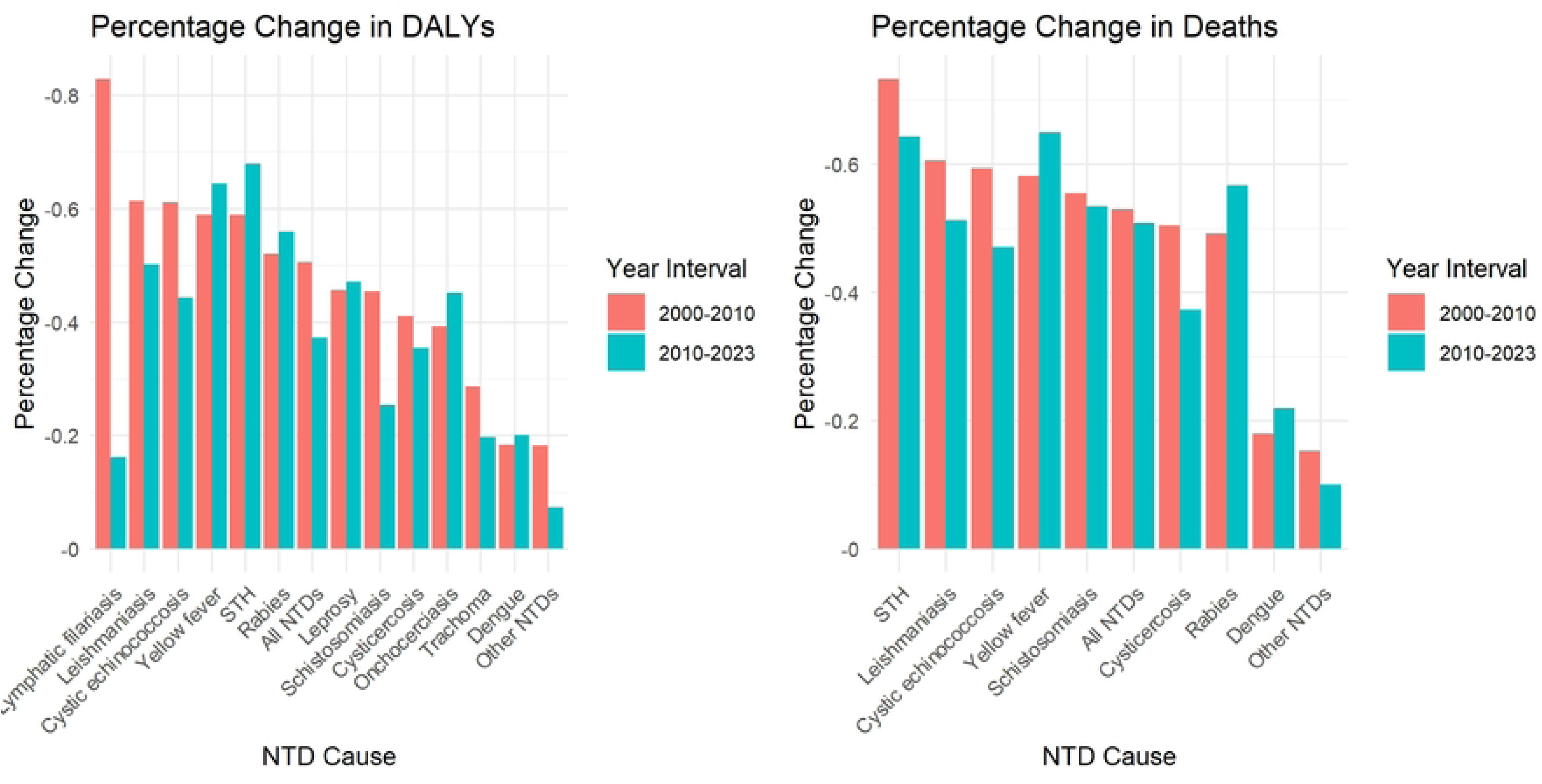
Percentage Change in Age-Standardized DALY and Mortality Rates for Major NTDs in Ethiopia, 2000–2010 and 2010–2023: GBD 2023 Estimates.

All major NTDs showed declining DALY trends nationally, although reductions varied by disease and period. Between 2010 and 2023, schistosomiasis DALYs declined by 25% (95% UI: 15%–38%), compared with a larger reduction of 45% (34%–55%) during 2000–2010. Similarly, reductions for trachoma slowed from 29% (24%–33%) in 2000–2010 to 20% (17%–23%) in 2010–2023. Lymphatic filariasis showed a marked deceleration, with reductions declining from 83% (75%–87%) to 16% (12%–20%) across the same periods (Figure 4).

At the subnational level, most regions showed declining ASDALY trends. However, schistosomiasis DALYs increased between 2010 and 2023 in Addis Ababa (25%) and Harari (13%), while trachoma DALYs increased by 9% in Oromia. Dengue fever showed increasing DALY burdens across most regions during the same period. In contrast, intestinal nematode infections, onchocerciasis, and yellow fever experienced larger reductions during 2010–2023 compared with 2000–2010 at both national and regional levels (Table 3).

**Table 3.** Trends in national and subnational age-standardized DALYs for NTDs: Percentage change for 1990–2023, 2000–2010, and 2010–2023.

| Region | year interval | ASDALY rate percentage change (95% UI), by specific NTDs |  |  |  |  |  |  |  |  |  |  |  |
| --- | --- | --- | --- | --- | --- | --- | --- | --- | --- | --- | --- | --- | --- |
|  |  | Schistosomiasis | Trachoma | Rabies | Other NTDs | Cysticercosis | STH | Yellow fever | L. filariasis | Leishmaniasis | Onchocerciasis | Dengue | Cystic echinococcosis |
| Addis Ababa | 1990-2023 | 0.39(-0.43,2.07) | -0.63(-0.67,-0.58) | -0.76(-0.94,0.47) | -0.35(-0.61,0.24) | -0.63(-0.78,-0.38) | -0.95(-0.97,-0.93) | -0.93(-0.98,-0.83) | -0.42(-0.47,-0.33) | -0.15(-0.49,0.54) | 0 | 0.35(-0.51,2.73) | -0.78(-0.87,-0.63) |
|  | 2000-2010 | 0.05(-0.50,1.69) | -0.37(-0.45,-0.29) | -0.3(-0.78,0.97) | -0.17(-0.41,0.26) | -0.38(-0.61,-0.03) | -0.85(-0.90,-0.77) | -0.39(-0.76,0.83) | -0.31(-0.39,-0.21) | -0.15(-0.24,0.01) | 0 | 0.2(-0.55,2.13) | -0.44(-0.65,-0.09) |
|  | 2010-2023 | 0.25(-0.49,1.74) | -0.31(-0.37,-0.24) | -0.35(-0.84,0.99) | -0.06(-0.39,0.57) | -0.32(-0.58,0.15) | -0.17(-0.50,0.41) | -0.6(-0.90,-0.01) | 0.05(-0.03,0.14) | -0.13(-0.27,0.07) | 0 | 0.19(-0.49,1.78) | -0.41(-0.63,-0.01) |
| Afar | 1990-2023 | -0.56(-0.72,-0.34) | -0.34(-0.38,-0.30) | -0.66(-0.95,0.96) | -0.14(-0.46,0.57) | -0.61(-0.74,-0.43) | -0.96(-0.98,-0.94) | -0.86(-0.95,-0.59) | -0.89(-0.93,-0.83) | -0.99(-0.99,-0.99) | 0 | 1.18(-0.64,8.64) | -0.84(-0.91,-0.70) |
|  | 2000-2010 | -0.24(-0.44,0.09) | -0.13(-0.17,-0.10) | -0.31(-0.80,1.70) | -0.06(-0.34,0.35) | -0.34(-0.51,-0.08) | -0.72(-0.79,-0.63) | -0.38(-0.76,0.64) | -0.81(-0.87,-0.73) | -0.47(-0.56,-0.34) | 0 | 0(-0.76,3.81) | -0.49(-0.73,-0.07) |
|  | 2010-2023 | -0.29(-0.53,-0.01) | -0.12(-0.17,-0.08) | -0.44(-0.90,1.56) | 0(-0.33,0.56) | -0.26(-0.47,0.06) | -0.56(-0.75,-0.33) | -0.57(-0.83,0.14) | -0.38(-0.45,-0.31) | -0.4(-0.53,-0.24) | 0 | 1.4(-0.50,9.91) | -0.43(-0.71,0.03) |
| Amhara | 1990-2023 | -0.63(-0.76,-0.46) | -0.46(-0.50,-0.41) | -0.86(-0.98,0.24) | -0.38(-0.66,0.45) | -0.72(-0.79,-0.62) | -0.91(-0.94,-0.87) | -0.93(-0.97,-0.86) | -0.89(-0.92,-0.83) | -0.99(-0.99,-0.98) | -0.74(-0.79,-0.67) | -0.4(-0.93,3.03) | -0.89(-0.92,-0.84) |
|  | 2000-2010 | -0.43(-0.56,-0.28) | -0.29(-0.35,-0.23) | -0.56(-0.89,1.22) | -0.26(-0.54,0.19) | -0.46(-0.59,-0.31) | -0.61(-0.69,-0.52) | -0.64(-0.82,-0.25) | -0.88(-0.92,-0.81) | -0.58(-0.65,-0.51) | -0.4(-0.46,-0.33) | -0.17(-0.82,3.12) | -0.65(-0.73,-0.52) |
|  | 2010-2023 | -0.28(-0.40,-0.17) | -0.23(-0.28,-0.19) | -0.62(-0.93,0.79) | -0.06(-0.50,0.76) | -0.38(-0.56,-0.15) | -0.51(-0.61,-0.39) | -0.63(-0.83,-0.34) | -0.07(-0.14,0.02) | -0.4(-0.49,-0.29) | -0.45(-0.52,-0.38) | -0.25(-0.89,6.06) | -0.45(-0.60,-0.21) |
| Benishangul-Gumuz | 1990-2023 | -0.59(-0.77,-0.27) | -0.4(-0.45,-0.36) | -0.81(-0.97,0.17) | -0.31(-0.59,0.51) | -0.58(-0.72,-0.35) | -0.98(-0.99,-0.97) | -0.92(-0.98,-0.33) | -0.94(-0.96,-0.91) | -0.99(-0.99,-0.99) | -0.73(-0.79,-0.66) | 1.07(-0.67,8.38) | -0.89(-0.94,-0.73) |
|  | 2000-2010 | -0.3(-0.62,0.22) | -0.15(-0.19,-0.12) | -0.41(-0.84,1.64) | -0.12(-0.43,0.39) | -0.26(-0.48,0.06) | -0.82(-0.88,-0.76) | -0.5(-0.87,2.68) | -0.61(-0.69,-0.52) | -0.63(-0.70,-0.55) | -0.38(-0.45,-0.31) | 0.05(-0.75,4.25) | -0.56(-0.80,0.06) |
|  | 2010-2023 | -0.3(-0.60,0.11) | -0.19(-0.25,-0.14) | -0.64(-0.93,0.68) | -0.15(-0.50,0.46) | -0.32(-0.52,0.00) | -0.79(-0.89,-0.65) | -0.73(-0.92,0.05) | -0.85(-0.88,-0.80) | -0.61(-0.68,-0.52) | -0.46(-0.53,-0.39) | 1.02(-0.57,8.84) | -0.56(-0.76,-0.16) |
| Dire Dawa | 1990-2023 | -0.73(-0.83,-0.59) | -0.74(-0.78,-0.70) | -0.88(-0.98,-0.14) | -0.34(-0.56,0.16) | -0.66(-0.79,-0.42) | -0.94(-0.97,-0.88) | -0.92(-1.00,611.73) | -0.87(-0.91,-0.80) | 0 | 0 | 0.56(-0.66,5.24) | -0.86(-0.94,-0.59) |
|  | 2000-2010 | -0.44(-0.65,-0.17) | -0.47(-0.52,-0.42) | -0.52(-0.85,0.88) | -0.16(-0.40,0.13) | -0.35(-0.59,-0.04) | -0.69(-0.88,-0.16) | -0.48(-1.00,1.00) | -0.75(-0.80,-0.66) | 0 | 0 | -0.04(-0.74,3.40) | -0.55(-0.81,0.17) |
|  | 2010-2023 | -0.42(-0.56,-0.25) | -0.22(-0.27,-0.16) | -0.65(-0.93,0.62) | -0.1(-0.38,0.39) | -0.35(-0.62,0.03) | -0.51(-0.80,0.25) | 0 | 0.01(-0.05,0.09) | 0 | 0 | 0.66(-0.56,5.34) | -0.52(-0.74,0.00) |
| Gambela | 1990-2023 | -0.66(-0.81,-0.24) | -0.66(-0.71,-0.62) | -0.89(-0.98,-0.30) | -0.35(-0.63,0.26) | -0.69(-0.80,-0.47) | -0.95(-0.98,-0.92) | -0.95(-1.00,1.00) | -0.77(-0.83,-0.67) | 0 | -0.74(-0.80,-0.68) | 0.82(-0.68,6.26) | -0.88(-0.94,-0.64) |
|  | 2000-2010 | -0.41(-0.69,0.18) | -0.34(-0.40,-0.27) | -0.52(-0.86,1.23) | -0.19(-0.47,0.20) | -0.36(-0.60,0.00) | -0.72(-0.84,-0.56) | -0.61(-1.00,1.00) | 0.2(0.12,0.28) | 0 | -0.4(-0.46,-0.34) | -0.12(-0.80,3.31) | -0.6(-0.82,-0.03) |
|  | 2010-2023 | -0.36(-0.65,0.11) | -0.32(-0.37,-0.28) | -0.64(-0.93,0.67) | -0.11(-0.48,0.52) | -0.4(-0.62,-0.03) | -0.44(-0.69,0.04) | -0.69(-1.00,320.20) | -0.8(-0.86,-0.73) | 0 | -0.46(-0.53,-0.40) | 1.05(-0.52,8.06) | -0.48(-0.70,0.03) |
| Harari | 1990-2023 | 0.06(-0.56,1.63) | -0.7(-0.76,-0.64) | -0.83(-0.97,0.07) | -0.32(-0.57,0.27) | -0.66(-0.80,-0.49) | -0.96(-0.98,-0.85) | 0 | -0.81(-0.85,-0.74) | 0 | 0 | 0.85(-0.67,7.13) | -0.82(-0.91,-0.47) |
|  | 2000-2010 | 0.01(-0.51,1.21) | -0.43(-0.50,-0.36) | -0.36(-0.84,1.25) | -0.16(-0.40,0.25) | -0.32(-0.55,0.06) | -0.69(-0.88,-0.24) | 0 | -0.64(-0.70,-0.58) | 0 | 0 | 0.02(-0.75,4.45) | -0.48(-0.79,0.31) |
|  | 2010-2023 | 0.13(-0.58,1.67) | -0.26(-0.30,-0.21) | -0.62(-0.93,0.82) | -0.06(-0.38,0.58) | -0.35(-0.61,-0.01) | -0.57(-0.80,0.13) | 0 | -0.01(-0.08,0.07) | 0 | 0 | 0.95(-0.55,8.48) | -0.43(-0.69,0.27) |
| **Oromia** | 1990-2023 | -0.73(-0.83,-0.59) | -0.38(-0.50,-0.18) | -0.82(-0.98,0.54) | -0.38(-0.62,0.07) | -0.73(-0.81,-0.65) | -0.96(-0.98,-0.94) | -0.92(-0.96,-0.83) | -0.86(-0.90,-0.79) | -0.85(-0.92,-0.69) | -0.73(-0.79,-0.66) | 0.47(-0.73,5.86) | -0.92(-0.94,-0.88) |
|  | 2000-2010 | -0.42(-0.59,-0.24) | -0.3(-0.39,-0.18) | -0.52(-0.89,1.25) | -0.17(-0.43,0.18) | -0.39(-0.53,-0.19) | -0.53(-0.62,-0.43) | -0.58(-0.80,-0.14) | -0.84(-0.88,-0.76) | -0.2(-0.33,-0.01) | -0.38(-0.45,-0.31) | -0.13(-0.78,3.57) | -0.64(-0.72,-0.52) |
|  | 2010-2023 | -0.18(-0.32,-0.06) | 0.09(0.01,0.18) | -0.52(-0.92,1.60) | -0.07(-0.39,0.42) | -0.34(-0.53,-0.10) | -0.79(-0.81,-0.75) | -0.63(-0.81,-0.21) | -0.03(-0.09,0.03) | -0.17(-0.33,0.05) | -0.45(-0.52,-0.39) | 1.06(-0.56,9.12) | -0.41(-0.55,-0.21) |
| **Sidama** | 1990-2023 | -0.68(-0.79,-0.51) | -0.69(-0.74,-0.64) | -0.8(-0.97,0.25) | -0.33(-0.67,0.50) | -0.64(-0.76,-0.44) | -0.91(-0.93,-0.88) | -0.9(-0.96,-0.76) | 0 | -0.99(-0.99,-0.99) | 0 | 1.4(-0.62,9.40) | -0.82(-0.90,-0.67) |
|  | 2000-2010 | -0.5(-0.60,-0.37) | -0.37(-0.43,-0.31) | -0.48(-0.84,0.83) | -0.17(-0.51,0.43) | -0.34(-0.55,-0.06) | -0.73(-0.78,-0.66) | -0.52(-0.78,-0.04) | 0 | -0.59(-0.65,-0.50) | 0 | 0.04(-0.76,3.90) | -0.51(-0.73,-0.19) |
|  | 2010-2023 | -0.25(-0.48,-0.02) | -0.39(-0.45,-0.34) | -0.61(-0.93,0.65) | -0.13(-0.53,0.63) | -0.36(-0.61,-0.04) | -0.54(-0.65,-0.39) | -0.64(-0.83,-0.18) | 0 | -0.52(-0.61,-0.40) | 0 | 1.04(-0.55,8.07) | -0.46(-0.68,-0.04) |
| **SNNP** | 1990-2023 | -0.75(-0.84,-0.60) | -0.65(-0.71,-0.59) | -0.8(-0.98,0.37) | -0.34(-0.64,0.28) | -0.72(-0.82,-0.57) | -0.9(-0.93,-0.87) | -0.94(-0.97,-0.88) | -0.86(-0.90,-0.78) | -0.99(-1.00,-0.99) | 0 | 0.84(-0.70,7.18) | -0.89(-0.93,-0.83) |
|  | 2000-2010 | -0.54(-0.64,-0.42) | -0.25(-0.32,-0.19) | -0.52(-0.86,0.95) | -0.17(-0.47,0.25) | -0.44(-0.60,-0.26) | -0.58(-0.67,-0.47) | -0.66(-0.84,-0.26) | -0.8(-0.86,-0.72) | -0.68(-0.73,-0.62) | 0 | -0.17(-0.80,3.13) | -0.63(-0.74,-0.49) |
|  | 2010-2023 | -0.34(-0.51,-0.12) | -0.45(-0.52,-0.38) | -0.53(-0.92,1.07) | -0.13(-0.48,0.48) | -0.38(-0.56,-0.15) | -0.69(-0.76,-0.60) | -0.73(-0.88,-0.43) | -0.18(-0.23,-0.13) | -0.67(-0.72,-0.58) | 0 | 1.23(-0.53,9.53) | -0.41(-0.61,-0.10) |
| **Somali** | 1990-2023 | -0.52(-0.68,-0.27) | -0.16(-0.22,-0.11) | -0.75(-0.97,0.42) | -0.14(-0.40,0.36) | -0.54(-0.69,-0.32) | -0.89(-0.92,-0.85) | -0.85(-0.93,-0.71) | -0.87(-0.91,-0.80) | -0.99(-0.99,-0.99) | 0 | 0.8(-0.69,6.79) | -0.83(-0.89,-0.71) |
|  | 2000-2010 | -0.32(-0.54,-0.02) | -0.08(-0.13,-0.02) | -0.42(-0.83,1.20) | -0.07(-0.34,0.24) | -0.26(-0.46,-0.01) | -0.58(-0.66,-0.50) | -0.34(-0.70,0.60) | -0.85(-0.90,-0.77) | -0.56(-0.63,-0.46) | 0 | -0.09(-0.77,3.52) | -0.49(-0.68,-0.23) |
|  | 2010-2023 | -0.16(-0.42,0.16) | -0.1(-0.15,-0.06) | -0.49(-0.92,1.45) | -0.01(-0.27,0.36) | -0.27(-0.53,0.05) | -0.54(-0.65,-0.40) | -0.6(-0.83,-0.20) | -0.21(-0.26,-0.15) | -0.53(-0.62,-0.42) | 0 | 1.08(-0.55,8.68) | -0.47(-0.65,-0.13) |
| **South West** | 1990-2023 | -0.81(-0.89,-0.71) | -0.7(-0.75,-0.66) | -0.83(-0.97,0.05) | -0.37(-0.68,0.58) | -0.71(-0.82,-0.51) | -0.9(-0.94,-0.85) | -0.92(-0.96,-0.82) | 0 | -0.99(-0.99,-0.99) | -0.73(-0.79,-0.67) | 0.9(-0.67,6.87) | -0.87(-0.93,-0.73) |
|  | 2000-2010 | -0.59(-0.70,-0.46) | -0.38(-0.44,-0.32) | -0.53(-0.86,0.60) | -0.21(-0.55,0.35) | -0.41(-0.61,-0.11) | -0.69(-0.77,-0.57) | -0.52(-0.79,0.00) | 0 | -0.59(-0.66,-0.50) | -0.39(-0.45,-0.32) | -0.05(-0.79,3.54) | -0.58(-0.78,-0.27) |
|  | 2010-2023 | -0.37(-0.57,-0.14) | -0.39(-0.45,-0.34) | -0.65(-0.94,0.46) | -0.14(-0.53,0.70) | -0.4(-0.61,-0.06) | -0.47(-0.63,-0.25) | -0.6(-0.81,-0.12) | 0 | -0.45(-0.55,-0.32) | -0.45(-0.52,-0.39) | 0.7(-0.61,6.64) | -0.53(-0.75,-0.10) |
| **Tigray** | 1990-2023 | -0.58(-0.72,-0.41) | -0.58(-0.61,-0.54) | -0.88(-0.98,-0.30) | -0.3(-0.59,0.37) | -0.7(-0.80,-0.56) | -0.98(-0.99,-0.97) | -0.91(-0.96,-0.81) | -0.83(-0.87,-0.77) | -0.99(-0.99,-0.98) | (0.00,0.00) | 1.09(-0.64,8.41) | -0.85(-0.90,-0.75) |
|  | 2000-2010 | -0.38(-0.53,-0.23) | -0.41(-0.46,-0.35) | -0.54(-0.87,0.79) | -0.23(-0.47,0.16) | -0.41(-0.59,-0.19) | -0.82(-0.86,-0.76) | -0.6(-0.80,-0.22) | -0.81(-0.85,-0.75) | -0.6(-0.66,-0.51) | (0.00,0.00) | -0.07(-0.78,3.66) | -0.59(-0.76,-0.32) |
|  | 2010-2023 | -0.16(-0.32,-0.03) | -0.19(-0.25,-0.14) | -0.71(-0.95,0.33) | -0.08(-0.44,0.57) | -0.38(-0.57,-0.07) | -0.64(-0.78,-0.48) | -0.66(-0.82,-0.30) | 0(-0.07,0.07) | -0.46(-0.55,-0.37) | (0.00,0.00) | 0.92(-0.58,9.51) | -0.45(-0.64,-0.09) |
| **Ethiopia** | 1990-2023 | -0.7(-0.80,-0.55) | -0.49(-0.54,-0.46) | -0.82(-0.98,0.25) | -0.34(-0.60,0.23) | -0.71(-0.77,-0.62) | -0.94(-0.96,-0.91) | -0.92(-0.95,-0.87) | -0.87(-0.90,-0.80) | -0.99(-0.99,-0.99) | -0.72(-0.78,-0.65) | -0.43(-0.93,2.55) | -0.89(-0.93,-0.84) |
|  | 2000-2010 | -0.45(-0.55,-0.34) | -0.29(-0.33,-0.24) | -0.52(-0.88,1.01) | -0.18(-0.46,0.19) | -0.41(-0.51,-0.28) | -0.59(-0.66,-0.52) | -0.59(-0.73,-0.41) | -0.83(-0.87,-0.75) | -0.61(-0.66,-0.55) | -0.39(-0.46,-0.32) | -0.18(-0.82,2.83) | -0.61(-0.71,-0.45) |
|  | 2010-2023 | -0.25(-0.38,-0.15) | -0.2(-0.23,-0.17) | -0.56(-0.92,1.05) | -0.07(-0.41,0.46) | -0.35(-0.47,-0.21) | -0.68(-0.72,-0.62) | -0.64(-0.75,-0.51) | -0.16(-0.20,-0.12) | -0.5(-0.58,-0.43) | -0.45(-0.52,-0.39) | -0.2(-0.84,5.25) | -0.44(-0.60,-0.21) |

STH-soil transmitted helminths; Othre NTDs-relapsing fevers, Lyme disease, typhus, arboviral and viral hemorrhagic fevers & other helminths; SNNP-Southern Nation Nationalities and Peoples region

Over the same period, the national ASMR for all NTDs declined from 72.3 per 100,000 in 1990 to 5.98 per 100,000 in 2023, corresponding to an overall reduction of 91.6%. Reductions were comparable across the two most recent decades, with declines of 53.3% during 2000–2010 and 50.4% during 2010–2023 (Figure 4). While national mortality trends remained largely stable across decades, schistosomiasis mortality increased in Addis Ababa (23%) and Harari (18%), and dengue fever mortality increased across most regions during 2010–2023 (Table 4).

**Table 4.**
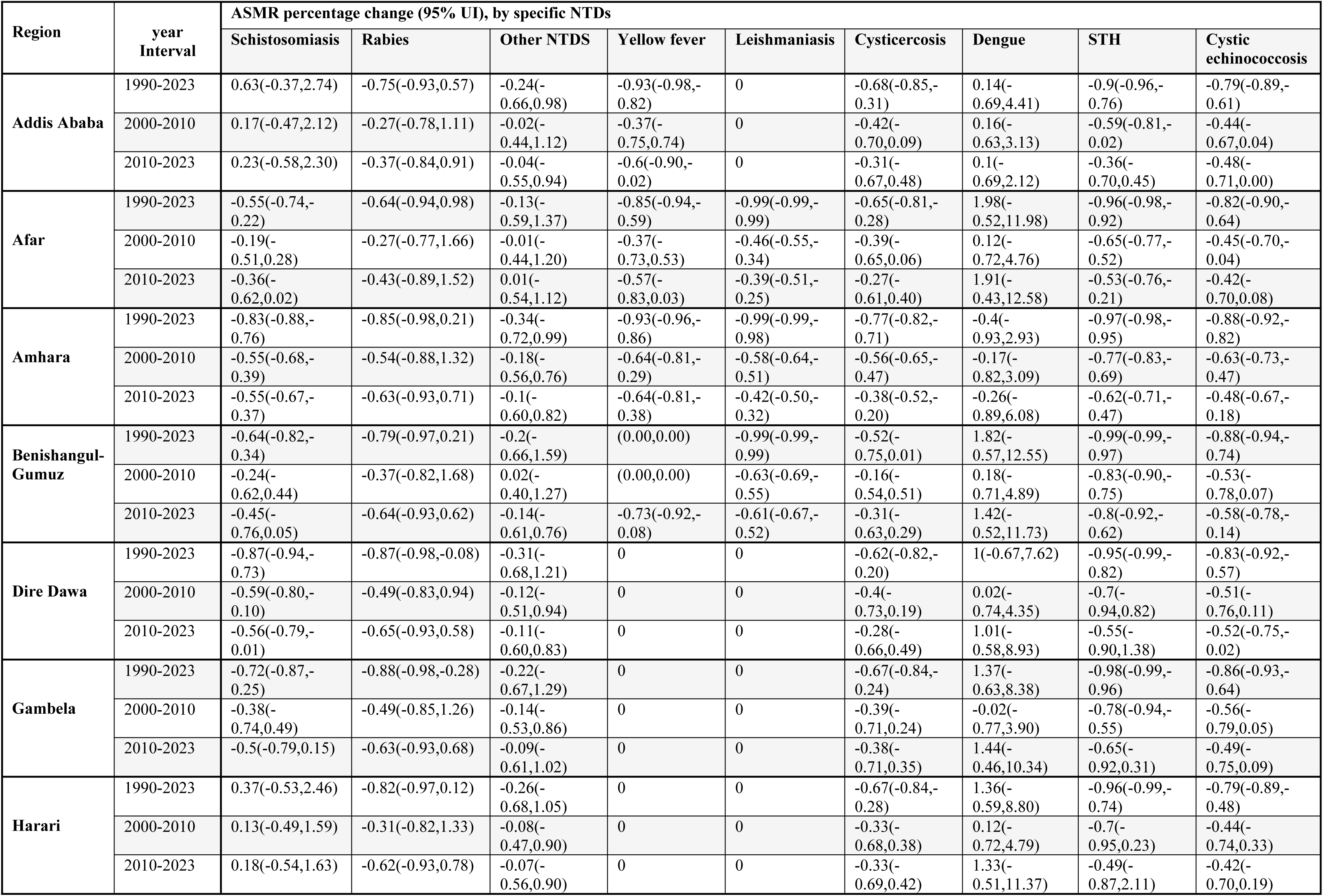
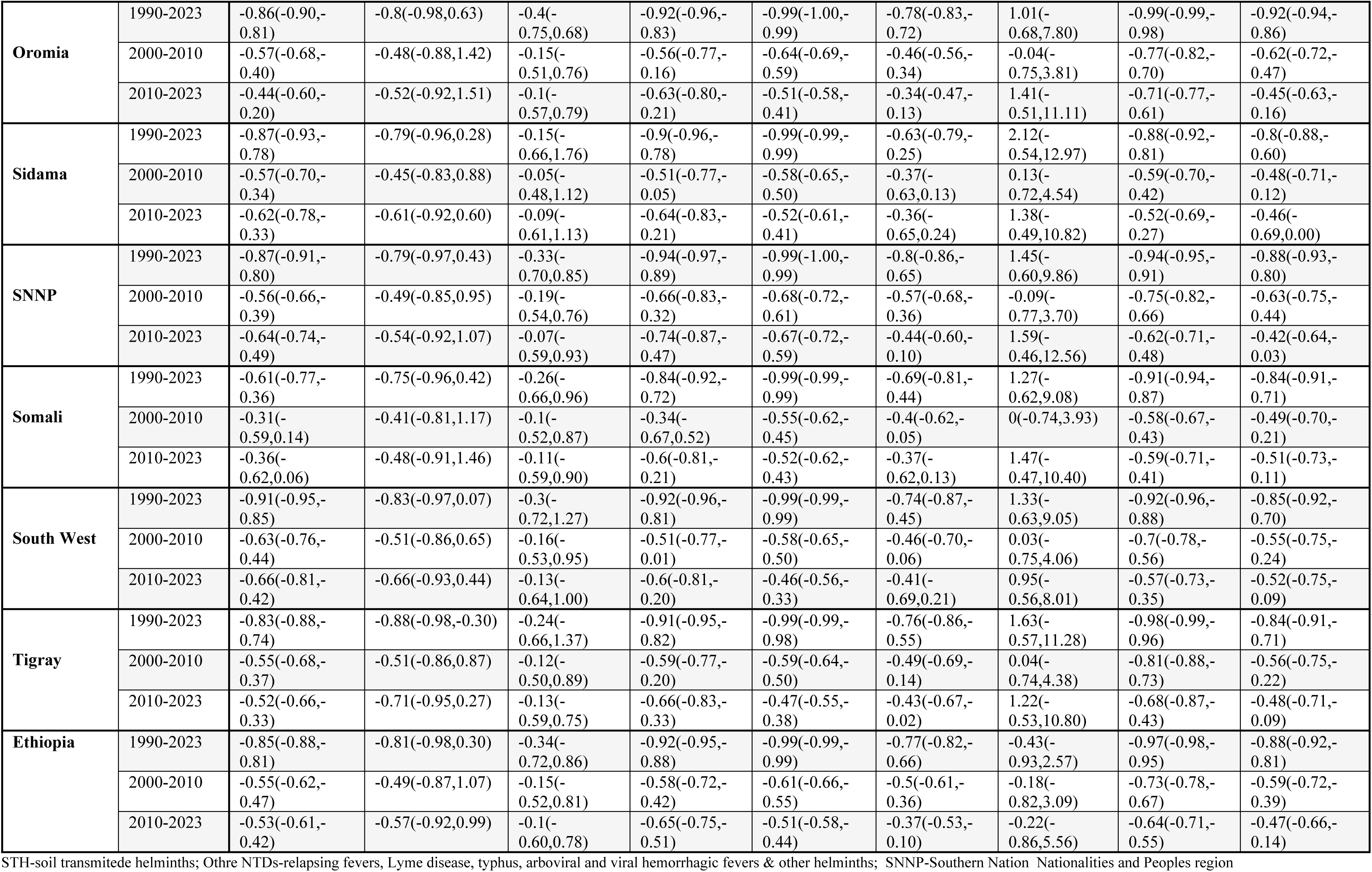
Trends in National and subnational age-standardized Mortality due to NTDs: Percentage change for 1990–2023, 2000–2010, and 2010–2023.

| Region | year Interval | ASMR percentage change (95% UI), by specific NTDs |  |  |  |  |  |  |  |  |
| --- | --- | --- | --- | --- | --- | --- | --- | --- | --- | --- |
|  |  | Schistosomiasis | Rabies | Other NTDS | Yellow fever | Leishmaniasis | Cysticercosis | Dengue | STH | Cystic echinococcosis |
| Addis Ababa | 1990-2023 | 0.63(-0.37,2.74) | -0.75(-0.93,0.57) | -0.24(-0.66,0.98) | -0.93(-0.98,-0.82) | 0 | -0.68(-0.85,-0.31) | 0.14(-0.69,4.41) | -0.9(-0.96,-0.76) | -0.79(-0.89,-0.61) |
|  | 2000-2010 | 0.17(-0.47,2.12) | -0.27(-0.78,1.11) | -0.02(-0.44,1.12) | -0.37(-0.75,0.74) | 0 | -0.42(-0.70,0.09) | 0.16(-0.63,3.13) | -0.59(-0.81,-0.02) | -0.44(-0.67,0.04) |
|  | 2010-2023 | 0.23(-0.58,2.30) | -0.37(-0.84,0.91) | -0.04(-0.55,0.94) | -0.6(-0.90,-0.02) | 0 | -0.31(-0.67,0.48) | 0.1(-0.69,2.12) | -0.36(-0.70,0.45) | -0.48(-0.71,0.00) |
| Afar | 1990-2023 | -0.55(-0.74,-0.22) | -0.64(-0.94,0.98) | -0.13(-0.59,1.37) | -0.85(-0.94,-0.59) | -0.99(-0.99,-0.99) | -0.65(-0.81,-0.28) | 1.98(-0.52,11.98) | -0.96(-0.98,-0.92) | -0.82(-0.90,-0.64) |
|  | 2000-2010 | -0.19(-0.51,0.28) | -0.27(-0.77,1.66) | -0.01(-0.44,1.20) | -0.37(-0.73,0.53) | -0.46(-0.55,-0.34) | -0.39(-0.65,0.06) | 0.12(-0.72,4.76) | -0.65(-0.77,-0.52) | -0.45(-0.70,-0.04) |
|  | 2010-2023 | -0.36(-0.62,0.02) | -0.43(-0.89,1.52) | 0.01(-0.54,1.12) | -0.57(-0.83,0.03) | -0.39(-0.51,-0.25) | -0.27(-0.61,0.40) | 1.91(-0.43,12.58) | -0.53(-0.76,-0.21) | -0.42(-0.70,0.08) |
| Amhara | 1990-2023 | -0.83(-0.88,-0.76) | -0.85(-0.98,0.21) | -0.34(-0.72,0.99) | -0.93(-0.96,-0.86) | -0.99(-0.99,-0.98) | -0.77(-0.82,-0.71) | -0.4(-0.93,2.93) | -0.97(-0.98,-0.95) | -0.88(-0.92,-0.82) |
|  | 2000-2010 | -0.55(-0.68,-0.39) | -0.54(-0.88,1.32) | -0.18(-0.56,0.76) | -0.64(-0.81,-0.29) | -0.58(-0.64,-0.51) | -0.56(-0.65,-0.47) | -0.17(-0.82,3.09) | -0.77(-0.83,-0.69) | -0.63(-0.73,-0.47) |
|  | 2010-2023 | -0.55(-0.67,-0.37) | -0.63(-0.93,0.71) | -0.1(-0.60,0.82) | -0.64(-0.81,-0.38) | -0.42(-0.50,-0.32) | -0.38(-0.52,-0.20) | -0.26(-0.89,6.08) | -0.62(-0.71,-0.47) | -0.48(-0.67,-0.18) |
| Benishangul-Gumuz | 1990-2023 | -0.64(-0.82,-0.34) | -0.79(-0.97,0.21) | -0.2(-0.66,1.59) | (0.00,0.00) | -0.99(-0.99,-0.99) | -0.52(-0.75,0.01) | 1.82(-0.57,12.55) | -0.99(-0.99,-0.97) | -0.88(-0.94,-0.74) |
|  | 2000-2010 | -0.24(-0.62,0.44) | -0.37(-0.82,1.68) | 0.02(-0.40,1.27) | (0.00,0.00) | -0.63(-0.69,-0.55) | -0.16(-0.54,0.51) | 0.18(-0.71,4.89) | -0.83(-0.90,-0.75) | -0.53(-0.78,0.07) |
|  | 2010-2023 | -0.45(-0.76,0.05) | -0.64(-0.93,0.62) | -0.14(-0.61,0.76) | -0.73(-0.92,-0.08) | -0.61(-0.67,-0.52) | -0.31(-0.63,0.29) | 1.42(-0.52,11.73) | -0.8(-0.92,-0.62) | -0.58(-0.78,-0.14) |
| Dire Dawa | 1990-2023 | -0.87(-0.94,-0.73) | -0.87(-0.98,-0.08) | -0.31(-0.68,1.21) | 0 | 0 | -0.62(-0.82,-0.20) | 1(-0.67,7.62) | -0.95(-0.99,-0.82) | -0.83(-0.92,-0.57) |
|  | 2000-2010 | -0.59(-0.80,-0.10) | -0.49(-0.83,0.94) | -0.12(-0.51,0.94) | 0 | 0 | -0.4(-0.73,0.19) | 0.02(-0.74,4.35) | -0.7(-0.94,0.82) | -0.51(-0.76,0.11) |
|  | 2010-2023 | -0.56(-0.79,-0.01) | -0.65(-0.93,0.58) | -0.11(-0.60,0.83) | 0 | 0 | -0.28(-0.66,0.49) | 1.01(-0.58,8.93) | -0.55(-0.90,1.38) | -0.52(-0.75,-0.02) |
| Gambela | 1990-2023 | -0.72(-0.87,-0.25) | -0.88(-0.98,-0.28) | -0.22(-0.67,1.29) | 0 | 0 | -0.67(-0.84,-0.24) | 1.37(-0.63,8.38) | -0.98(-0.99,-0.96) | -0.86(-0.93,-0.64) |
|  | 2000-2010 | -0.38(-0.74,0.49) | -0.49(-0.85,1.26) | -0.14(-0.53,0.86) | 0 | 0 | -0.39(-0.71,0.24) | -0.02(-0.77,3.90) | -0.78(-0.94,-0.55) | -0.56(-0.79,0.05) |
|  | 2010-2023 | -0.5(-0.79,0.15) | -0.63(-0.93,0.68) | -0.09(-0.61,1.02) | 0 | 0 | -0.38(-0.71,0.35) | 1.44(-0.46,10.34) | -0.65(-0.92,0.31) | -0.49(-0.75,0.09) |
| Harari | 1990-2023 | 0.37(-0.53,2.46) | -0.82(-0.97,0.12) | -0.26(-0.68,1.05) | 0 | 0 | -0.67(-0.84,-0.28) | 1.36(-0.59,8.80) | -0.96(-0.99,-0.74) | -0.79(-0.89,-0.48) |
|  | 2000-2010 | 0.13(-0.49,1.59) | -0.31(-0.82,1.33) | -0.08(-0.47,0.90) | 0 | 0 | -0.33(-0.68,0.38) | 0.12(-0.72,4.79) | -0.7(-0.95,0.23) | -0.44(-0.74,0.33) |
|  | 2010-2023 | 0.18(-0.54,1.63) | -0.62(-0.93,0.78) | -0.07(-0.56,0.90) | 0 | 0 | -0.33(-0.69,0.42) | 1.33(-0.51,11.37) | -0.49(-0.87,2.11) | -0.42(-0.70,0.19) |
| **Oromia** | 1990-2023 | -0.86(-0.90,-0.81) | -0.8(-0.98,0.63) | -0.4(-0.75,0.68) | -0.92(-0.96,-0.83) | -0.99(-1.00,-0.99) | -0.78(-0.83,-0.72) | 1.01(-0.68,7.80) | -0.99(-0.99,-0.98) | -0.92(-0.94,-0.86) |
|  | 2000-2010 | -0.57(-0.68,-0.40) | -0.48(-0.88,1.42) | -0.15(-0.51,0.76) | -0.56(-0.77,-0.16) | -0.64(-0.69,-0.59) | -0.46(-0.56,-0.34) | -0.04(-0.75,3.81) | -0.77(-0.82,-0.70) | -0.62(-0.72,-0.47) |
|  | 2010-2023 | -0.44(-0.60,-0.20) | -0.52(-0.92,1.51) | -0.1(-0.57,0.79) | -0.63(-0.80,-0.21) | -0.51(-0.58,-0.41) | -0.34(-0.47,-0.13) | 1.41(-0.51,11.11) | -0.71(-0.77,-0.61) | -0.45(-0.63,-0.16) |
| **Sidama** | 1990-2023 | -0.87(-0.93,-0.78) | -0.79(-0.96,0.28) | -0.15(-0.66,1.76) | -0.9(-0.96,-0.78) | -0.99(-0.99,-0.99) | -0.63(-0.79,-0.25) | 2.12(-0.54,12.97) | -0.88(-0.92,-0.81) | -0.8(-0.88,-0.60) |
|  | 2000-2010 | -0.57(-0.70,-0.34) | -0.45(-0.83,0.88) | -0.05(-0.48,1.12) | -0.51(-0.77,-0.05) | -0.58(-0.65,-0.50) | -0.37(-0.63,0.13) | 0.13(-0.72,4.54) | -0.59(-0.70,-0.42) | -0.48(-0.71,-0.12) |
|  | 2010-2023 | -0.62(-0.78,-0.33) | -0.61(-0.92,0.60) | -0.09(-0.61,1.13) | -0.64(-0.83,-0.21) | -0.52(-0.61,-0.41) | -0.36(-0.65,0.24) | 1.38(-0.49,10.82) | -0.52(-0.69,-0.27) | -0.46(-0.69,0.00) |
| **SNNP** | 1990-2023 | -0.87(-0.91,-0.80) | -0.79(-0.97,0.43) | -0.33(-0.70,0.85) | -0.94(-0.97,-0.89) | -0.99(-1.00,-0.99) | -0.8(-0.86,-0.65) | 1.45(-0.60,9.86) | -0.94(-0.95,-0.91) | -0.88(-0.93,-0.80) |
|  | 2000-2010 | -0.56(-0.66,-0.39) | -0.49(-0.85,0.95) | -0.19(-0.54,0.76) | -0.66(-0.83,-0.32) | -0.68(-0.72,-0.61) | -0.57(-0.68,-0.36) | -0.09(-0.77,3.70) | -0.75(-0.82,-0.66) | -0.63(-0.75,-0.44) |
|  | 2010-2023 | -0.64(-0.74,-0.49) | -0.54(-0.92,1.07) | -0.07(-0.59,0.93) | -0.74(-0.87,-0.47) | -0.67(-0.72,-0.59) | -0.44(-0.60,-0.10) | 1.59(-0.46,12.56) | -0.62(-0.71,-0.48) | -0.42(-0.64,-0.03) |
| **Somali** | 1990-2023 | -0.61(-0.77,-0.36) | -0.75(-0.96,0.42) | -0.26(-0.66,0.96) | -0.84(-0.92,-0.72) | -0.99(-0.99,-0.99) | -0.69(-0.81,-0.44) | 1.27(-0.62,9.08) | -0.91(-0.94,-0.87) | -0.84(-0.91,-0.71) |
|  | 2000-2010 | -0.31(-0.59,0.14) | -0.41(-0.81,1.17) | -0.1(-0.52,0.87) | -0.34(-0.67,0.52) | -0.55(-0.62,-0.45) | -0.4(-0.62,-0.05) | 0(-0.74,3.93) | -0.58(-0.67,-0.43) | -0.49(-0.70,-0.21) |
|  | 2010-2023 | -0.36(-0.62,0.06) | -0.48(-0.91,1.46) | -0.11(-0.59,0.90) | -0.6(-0.81,-0.21) | -0.52(-0.62,-0.43) | -0.37(-0.62,0.13) | 1.47(-0.47,10.40) | -0.59(-0.71,-0.41) | -0.51(-0.73,-0.11) |
| **South West** | 1990-2023 | -0.91(-0.95,-0.85) | -0.83(-0.97,0.07) | -0.3(-0.72,1.27) | -0.92(-0.96,-0.81) | -0.99(-0.99,-0.99) | -0.74(-0.87,-0.45) | 1.33(-0.63,9.05) | -0.92(-0.96,-0.88) | -0.85(-0.92,-0.70) |
|  | 2000-2010 | -0.63(-0.76,-0.44) | -0.51(-0.86,0.65) | -0.16(-0.53,0.95) | -0.51(-0.77,-0.01) | -0.58(-0.65,-0.50) | -0.46(-0.70,-0.06) | 0.03(-0.75,4.06) | -0.7(-0.78,-0.56) | -0.55(-0.75,-0.24) |
|  | 2010-2023 | -0.66(-0.81,-0.42) | -0.66(-0.93,0.44) | -0.13(-0.64,1.00) | -0.6(-0.81,-0.20) | -0.46(-0.56,-0.33) | -0.41(-0.69,0.21) | 0.95(-0.56,8.01) | -0.57(-0.73,-0.35) | -0.52(-0.75,-0.09) |
| **Tigray** | 1990-2023 | -0.83(-0.88,-0.74) | -0.88(-0.98,-0.30) | -0.24(-0.66,1.37) | -0.91(-0.95,-0.82) | -0.99(-0.99,-0.98) | -0.76(-0.86,-0.55) | 1.63(-0.57,11.28) | -0.98(-0.99,-0.96) | -0.84(-0.91,-0.71) |
|  | 2000-2010 | -0.55(-0.68,-0.37) | -0.51(-0.86,0.87) | -0.12(-0.50,0.89) | -0.59(-0.77,-0.20) | -0.59(-0.64,-0.50) | -0.49(-0.69,-0.14) | 0.04(-0.74,4.38) | -0.81(-0.88,-0.73) | -0.56(-0.75,-0.22) |
|  | 2010-2023 | -0.52(-0.66,-0.33) | -0.71(-0.95,0.27) | -0.13(-0.59,0.75) | -0.66(-0.83,-0.33) | -0.47(-0.55,-0.38) | -0.43(-0.67,-0.02) | 1.22(-0.53,10.80) | -0.68(-0.87,-0.43) | -0.48(-0.71,-0.09) |
| **Ethiopia** | 1990-2023 | -0.85(-0.88,-0.81) | -0.81(-0.98,0.30) | -0.34(-0.72,0.86) | -0.92(-0.95,-0.88) | -0.99(-0.99,-0.99) | -0.77(-0.82,-0.66) | -0.43(-0.93,2.57) | -0.97(-0.98,-0.95) | -0.88(-0.92,-0.81) |
|  | 2000-2010 | -0.55(-0.62,-0.47) | -0.49(-0.87,1.07) | -0.15(-0.52,0.81) | -0.58(-0.72,-0.42) | -0.61(-0.66,-0.55) | -0.5(-0.61,-0.36) | -0.18(-0.82,3.09) | -0.73(-0.78,-0.67) | -0.59(-0.72,-0.39) |
|  | 2010-2023 | -0.53(-0.61,-0.42) | -0.57(-0.92,0.99) | -0.1(-0.60,0.78) | -0.65(-0.75,-0.51) | -0.51(-0.58,-0.44) | -0.37(-0.53,-0.10) | -0.22(-0.86,5.56) | -0.64(-0.71,-0.55) | -0.47(-0.66,-0.14) |

STH-soil transmitted helminths; Othre NTDs-relapsing fevers, Lyme disease, typhus, arboviral and viral hemorrhagic fevers & other helminths; SNNP-Southern Nation Nationalities and Peoples region

## Discussions

This study comprehensively analyzed national and subnational GBD estimates to assess the DALYs and mortality burden of neglected tropical diseases in Ethiopia. Because several common NTDs including trachoma, lymphatic filariasis, onchocerciasis, and STH rarely result in death, DALYs remain the most appropriate measure of their public health impact. In 2023, Ethiopia was affected by 13 NTDs nationally, with all regions and city administrations estimated to be endemic to 10-13 diseases, the persistent and widespread nature of NTDs as public health challenge. Consistent with previous studies and national mapping surveys, schistosomiasis, trachoma, rabies, intestinal nematodes, and onchocerciasis were the leading contributors to NTD-related DALYs, while schistosomiasis, rabies, yellow fever, and leishmaniasis accounted for the majority of NTD-related deaths at both national and subnational levels. These findings align with national mapping reports which documented the widespread endemicity of trachoma, soil-transmitted helminths, schistosomiasis, and onchocerciasis affecting nearly three-quarters of the population and up to 798 districts(14,18,22).

This study highlighted that subnational disparities were evident, with regions such as Afar, Amhara, Tigray, South west and SNNPR bearing a disproportionately higher NTD burden compared with other regions. These variations may be attributed to differences in ecological suitability for disease transmission, vector distribution, access to water and sanitation, socioeconomic status, and health service availability and access. The low burden observed in Addis Ababa and Dire Dawa likely reflects better access to healthcare and improved WASH coverage in urban settings. Similar patterns have been reported in earlier national and subnational studies, reinforcing the dominant contribution of these diseases to Ethiopia’s overall NTD burden(15,17,25).

Sex-specific patterns showed that the overall DALY burden was similar between males and females; however, specific diseases displayed notable differences. Rabies imposed a higher burden on females, whereas yellow fever predominantly affected males. Mortality patterns were consistent with these observations, highlighting the need for gender based intervention strategies. Age specific analyses revealed disease specific vulnerabilities. Schistosomiasis and rabies affected all age groups relatively evenly, while STH and other NTDs primarily impacted children under 15 years. Trachoma burden peaked among older adults (≥50 years), and yellow fever mainly affected young adults (15–35 years). Mortality mirrored these patterns, with schistosomiasis and other NTD deaths increasing with age, particularly among adults ≥60 years. These findings underscore the importance of age-targeted interventions like, school-based programs for childhood NTDs and adult-focused strategies for trachoma, rabies, and yellow fever.

Overall, the burden of NTDs has decreased substantially over the last three decades. From 1990 to 2023, Ethiopia achieved an 87.5% reduction in NTD-related DALYs and a 91.6% decline in mortality. Reductions were observed across all regions, indicating the widespread benefits of national intervention efforts. This progress coincides with intensified global and local actions following the global and subsequent implementation of Ethiopia’s national NTD strategic plan (20,22). Expanded access to mass drug administration, integrated vector management, intensified case management, and the SAFE strategy for trachoma are likely contributors to these gains (13,14,18). Additionally, socioeconomic improvements and expanded healthcare delivery may have indirectly supported reductions in exposure and disease severity.

Despite these achievements, the rate of progress has slowed in recent years. Between 2010 and 2023, the decline in DALYs for major NTDs particularly schistosomiasis, trachoma, and lymphatic filariasis was markedly lower than reductions achieved between 2000 and 2010. Furthermore, significant subnational increases were observed. DALYs due to schistosomiasis increased in Addis Ababa and Harari, trachoma rose in Oromia, and dengue fever showed rising burdens across multiple regions. These trends were mirrored in mortality patterns, with schistosomiasis and dengue fever showing regional increases between 2010 and 2023. Such stagnation and localized surges may reflect programmatic gaps, inadequate disease-specific interventions, poor WASH improvements in hotspots, climate and environmental changes favoring vector proliferation, the COVID-19 pandemic’s disruption of routine health services, and sociopolitical instability limiting intervention coverage(13,17).

The observed deceleration in progress poses a challenge to achieving Ethiopia’s national NTD targets. Current trends suggest that the country is unlikely to meet its planned reductions without intensified, targeted, and context-specific interventions. Strengthening disease-specific programs, prioritizing high-burden and stagnating regions, and reinforcing multisectoral collaboration particularly with WASH, environmental management, and veterinary public health will be critical to accelerating progress.

This study has limitations inherent to the use of GBD modeled estimates, which depend on the availability and quality of underlying data. In settings with limited or incomplete surveillance and reporting, subnational estimates may be subject to greater uncertainty. Strengthening routine health information systems, laboratory confirmation, and disease-specific surveillance will be essential to improve the precision of future estimates and guide targeted interventions.

## Conclusion

Neglected tropical diseases remain a significant public health challenge in Ethiopia, with substantial disability and mortality and marked subnational variation. Although major reductions in NTD burden have been achieved since 1990, progress has slowed in recent years, and some diseases show increasing trends in specific regions. At the current pace, national NTD reduction targets are unlikely to be met without intensified and targeted action. Age- and sex-specific patterns highlight the need for tailored interventions, including school-based programs for children, adult-focused strategies for older populations, and gender-sensitive approaches where appropriate. Sustained investment in core NTD interventions, strengthened surveillance systems, improved WASH infrastructure, and focused attention on high-burden and emerging hotspot regions are essential to accelerate progress toward Ethiopia’s NTD control and elimination goals.

## Data Availability

The data usde in this study is the GBD study data which is publicy avalable online and it is indicated in the manuscript.

## Acknowledgments

We, the authors, are grateful to the collaboration between National Data Management and Analytics Centre (NDMC) at Ethiopian Public Health Institutes, Institute of Health and Metric Evaluation (IHME) at University of Washington and GBD Collaborators Network of Experts for Ethiopia national and subnational burden of disease analysis.

